# Older more fit KL-VS heterozygotes have more favorable AD-relevant biomarker profiles

**DOI:** 10.1101/2025.02.27.25323056

**Authors:** Mackenzie Jarchow, Ira Driscoll, Brianne M. Breidenbach, Noah Cook, Catherine L. Gallagher, Sterling C. Johnson, Sanjay Asthana, Bruce P. Hermann, Mark A. Sager, Kaj Blennow, Henrik Zetterberg, Cynthia M. Carlsson, Gwendlyn Kollmorgen, Clara Quijano-Rubio, Dane B. Cook, Dena B. Dubal, Ozioma C. Okonkwo

**Author notes:** **Correspondence:** Ira (Frahmand) Driscoll, PhD, University of Wisconsin - Madison, J5/M192 Clinical Science Center, 600 Highland Avenue, Madison, WI 53792, Ozioma Okonkwo, PhD, University of Wisconsin, Madison, J5/156M, Clinical Science Center, 600 Highland Avenue, Madison, WI 53792.

## Abstract

**INTRODUCTION:** While hallmarked by the accumulation of β-amyloid plaques (Aβ) and neurofibrillary tangles (tau) in the brain, Alzheimer’s disease (AD) is a multifactorial disorder that involves additional pathological events, including neuroinflammation, neurodegeneration and synaptic dysfunction. AD-associated biomolecular changes seem to be attenuated in carriers of the functionally advantageous variant of the *KLOTHO* gene (KL-VS_HET_). Independently, better cardiorespiratory fitness (CRF) is associated with better health outcomes, both in general and specifically with regard to AD pathology. Here we investigate whether the relationships between CRF (peak oxygen consumption (VO_2peak_)) and cerebrospinal fluid (CSF) core AD biomarkers and those of neuroinflammation, neurodegeneration, and synaptic dysfunction differ for KL-VS_HET_ compared to non-carriers (KL-VS_NC_).

**METHODS:** The cohort, enriched for AD risk, consisted of cognitively unimpaired adults (N=136; Mean_AGE_(SD)=62.5(6.7)) from the Wisconsin Registry for Alzheimer’s Prevention and the Wisconsin Alzheimer’s Disease Research Center. Covariate-adjusted (age, sex, parental AD history, *APOE*4+ status, and age difference between CSF sampling and exercise test) linear models examined the interaction between VO_2peak_ and *KLOTHO* genotype on core AD biomarker levels in CSF [phosphorylated tau 181 (pTau_181_), Aβ_42_/Aβ_40_, pTau_181_/Aβ_42_]. Analyses were repeated for CSF biomarkers of <u>neurodegeneration</u> [total tau (tTau), α-synuclein (α-syn), neurofilament light polypeptide (NfL)], <u>synaptic dysfunction</u> [neurogranin (Ng)], and <u>neuroinflammation</u> [glial fibrillary acidic protein (GFAP), soluble triggering receptor expressed in myeloid cells (sTREM2), chitinase-3-like protein 1 (YKL-40), interleukin 6 (IL-6), S100 calcium-binding protein B (S100B)].

**RESULTS:** The interaction between VO_2peak_ and KL-VS_HET_ was significant for **tTau** (*P*=0.05), **pTau_181_** (*P*=0.03), **Ng** (*P*=0.02), **sTREM2** (*P*=0.03), and **YKL-40** (*P*=0.03), such that lower levels of each biomarker were observed for KL-VS_HET_ who were more fit. No significant KL-VSxVO_2peak_ interactions were observed for Aβ_42_/Aβ_40_, pTau_181_/Aβ_42_, α-syn, NfL, GFAP, IL-6 or S100B (all *P*s>0.09).

**CONCLUSIONS:** We report a synergistic relationship between KL-VS_HET_ and CRF with regard to pTau_181_, tTau, Ng, sTREM2 and YKL-40, suggesting a protective role for both KL-VS_HET_ and better cardiovascular fitness against unfavorable AD-related changes. Their potentially shared biological mechanisms will require future investigations.

**Research in Context:** *Systematic Review:* PubMed literature review suggests that both *KLOTHO* KL-VS genotype and cardiorespiratory fitness (CRF) are associated with pathophysiological processes related to Alzheimer’s Disease (AD). Both KL-VS heterozygotes (KL-VS_HET_) and those with higher CRF fare better when faced with age-related biomolecular changes of relevance to AD. The present study investigates whether the relationships between CRF and cerebrospinal fluid biomarkers (CSF) of core AD neuropathology, neuroinflammation, neurodegeneration, and synaptic dysfunction differ for KL-VS_HET_ compared to non-carriers.

*Interpretation:* Our findings suggest a synergistic relationship between KL-VS_HET_ and higher CRF against core AD pathology along a range of unfavorable biomolecular changes implicated in this multifactorial disease. This supports the idea that CRF may interact with genetic factors to confer resilience against a multitude of adverse AD-associated processes.

*Future Directions:* Future studies should examine longitudinal changes in CSF biomarkers to determine whether maintaining or improving CRF over time enhances AD resilience in KL-VS_HET_.

## 2 Introduction

Alzheimer’s disease (AD), the most prevalent type of dementia, has garnered increasing attention as a pressing public health concern, particularly in light of the growing elderly population [1]. Given that age is the most significant risk factor for AD, this demographic shift is expected to lead to a marked rise in the number of individuals afflicted by this neurodegenerative disease [1]. Characterized by distinct pathophysiological hallmarks, the accumulation of beta-amyloid (Aβ) plaques and neurofibrillary tangles (tau) in the brain, AD causes irreparable cognitive and functional impairments [2]. This growing burden—affecting over 55 million people globally—highlights the critical need for advancing research efforts to develop preventative measures and therapeutic strategies to combat AD [1]. There is a growing interest in research surrounding modifiable and non-modifiable factors that might mitigate AD risk. Two such factors, a functionally advantageous *KLOTHO* KL-VS genotype (non-modifiable) and cardiorespiratory fitness (CRF; modifiable), are of particular interest as they are both associated with various favorable outcomes related to AD [3–12].

*KLOTHO* is considered an anti-aging and longevity gene that encodes klotho, a transmembrane protein responsible for regulating various aging processes in mammals [13, 14] . In humans, two genetic variants of *KLOTHO*, rs9536314 and rs9527025, combine to form a functional haplotype known as KL-VS [15]. The functionally advantageous KL-VS genotype (KL-VS_HET_) is associated with higher circulating klotho protein levels and more favorable outcomes related to cardiovascular health, renal sufficiency, and cognitive function [14–17] within the context of aging. More recent literature examining *KLOTHO* in relation to AD suggests that KL-VS_HET_ is associated with lower Aβ aggregation [11], tau burden [3, 4] and AD risk in apolipoprotein E (*APOE*) ε4 carriers [18]. Additionally, KL-VS_HET_’s protective properties seem to extend to deleterious age-related changes in synaptic integrity, neurodegeneration, and neuroinflammatory responses [12]. This apparent broad neuroprotection suggests a critical role for KL-VS_HET_ in safeguarding the brain from AD-associated biomolecular changes and related pathological events [3, 4, 11, 12].

CRF, a widely recognized index of habitual physical activity, is associated with reduced risk for AD-related dementia mortality [19], slower rates of gray matter atrophy in AD-relevant brain regions [5, 10], attenuation of white matter hyperintensities [20], and preservation of hippocampal volume with age [21]. Furthermore, emerging evidence suggests that physical exercise positively impacts not only cognition [5, 6, 10, 21] but also the underlying disease mechanisms, including Aβ deposition [7, 22, 23] and tau burden [24]. Additionally, higher CRF is also associated with less neuroinflammation [25] and better neuroplasticity [26]. The literature highlights the potential of CRF as a therapeutic strategy to address various pathological mechanisms of AD.

While both KL-VS_HET_ and CRF seem to confer resilience against deleterious AD-associated changes, there is a notable absence of research that concurrently examines the synergy between these two factors with respect to their combined impact on biomolecular changes associated with AD. Moreover, existent interventions largely focus on mitigating Aβ and tau burden, even though AD is a multifactorial disease involving additional pathological processes, such as neuroinflammation, neurodegeneration, and synaptic dysfunction [27]. Hence, the objective of the current study is to investigate whether the relationships between CRF, indexed here by VO_2peak_, and cerebrospinal fluid (CSF) biomarkers of core AD pathology, neurodegeneration, synaptic dysfunction, and neuroinflammation differ for *KLOTHO* KL-VS non-carriers (KL-VS_NC_) and KL-VS_HET_ in an older, cognitively unimpaired cohort enriched for AD risk. We hypothesize that KL-VS_HET_ individuals with higher CRF will have a more favorable profile of investigated CSF biomarkers compared to KL-VS_NC_.

## 3 Methods

### 3.1 Participants

The current sample is comprised of 136 (Mean_AGE_(SD) = 62.5(6.7); 64% female) cognitively unimpaired, middle-aged and older adults from the Wisconsin Registry for Alzheimer’s Prevention (WRAP) [28] and Wisconsin Alzheimer’s Disease Research Center (WADRC) [11] who were genotyped for *APOE* and *KLOTHO*, underwent CSF sampling, and had available VO_2peak_ data. The cohort was enriched for parental history of AD at enrollment, and subsequently has a higher prevalence of *APOE* ε4 allele carriers (*APOE*4+) than what is observed in the general population. Cognitive normalcy was determined by a standardized and multidisciplinary consensus based on performance on a comprehensive battery of neuropsychological tests, lack of functional impairment, and absence of neurologic/psychiatric conditions that might impair cognition [11, 28]. All study procedures were approved by the Institutional Review Board at the University of Wisconsin and each participant provided written informed consent before taking part.

### 3.2 Genotyping

Using the PUREGENE DNA Isolation Kit (Gentra Systems, Inc., Minneapolis, MN), DNA was isolated from whole-blood samples. Ultraviolet spectrophotometry (DU 530 Spectrophotometer, Beckman Coulter, Fullerton, CA) was used to measure DNA concentrations. Genotyping for *APOE* (rs429358 and rs7412) and *KLOTHO* (rs9536314 for F352V and rs9527025 for C370S) was done by LGC Genomics (Beverly, MA) via competitive allele-specific PCR-based KASP genotyping assays. Quality control procedures have been previously published [11] and are considered acceptable. KL-VS homozygosity, a rare genotype associated with lower klotho levels [16], was excluded from analyses due to sparse sample size (N=6).

### 3.3 CSF Assessment

After a 12-hour fast, a lumbar puncture was conducted at L3-4 or L4-5 using a drip method and/or gentle extraction technique into polypropylene syringes using a Sprotte 24- or 25-gauge spinal needle. A thin needle was used to inject 1% lidocaine as a local anesthetic prior to the insertion of the Sprotte spinal needle. 22 milliliters of CSF from each sample were pooled, gently mixed, and centrifuged at 2,000 g for 10 minutes. Supernatants were kept at 80°C and frozen in 0.5 milliliters aliquots in polypropylene tubes.

Samples were analyzed for phosphorylated tau (pTau_181_), Aβ_40,_ and Aβ_42_ to obtain biomarkers of core **AD pathology** (pTau_181_, Aβ_42_/Aβ_40,_ and pTau_181_/Aβ_42_), **neurodegeneration** [total tau (tTau), α-synuclein (α-syn), neurofilament light polypeptide (NfL)], **synaptic dysfunction** [neurogranin (Ng)], and **neuroinflammation** [glial fibrillary acidic protein (GFAP), soluble triggering receptor expressed in myeloid cells 2 (sTREM2), chitinase-3-like protein 1 (YKL-40), interleukin 6 (IL-6), S100 calcium-binding protein B (S100B)]) using the NeuroToolKit (NTK; Roche Diagnostics International Ltd, Rotkruez, Switzerland). The NTK is a panel of exploratory prototype assays designed to robustly evaluate established AD biomarkers (Aβ and tau) as well as emerging markers. This toolkit enables a comprehensive characterization of AD pathology as well as a panel of synaptic, axonal, and glial biomarkers, providing enhanced insights into the disease’s pathophysiological processes [29].

### 3.4 Cardiorespiratory Fitness (VO_2peak_) Assessment

Details of VO_2peak_assessment have been previously published [6, 20]. To assess VO_2peak_, the gold standard for direct assessment of CRF [30], participants completed a maximal graded exercise test administered by a certified exercise physiologist using standard operating procedures defined by American College of Sports Medicine guidelines. Participants underwent medical screening, including a resting ECG, to ensure safety before testing. Heart rate and rhythm were monitored continuously alongside oxygen uptake (VO), carbon dioxide production, and other respiratory metrics using a calibrated metabolic cart (TrueOne® 2400, Parvomedics). Participants walked at a self-selected speed, with treadmill incline increasing 2.5% every two minutes until volitional exhaustion. Peak effort was defined by at least two of the following criteria: (1) respiratory exchange ratio greater than or equal to 1.1, (2) achievement of 90% of age-predicted maximum heart rate, (3) perceived exertion greater than or equal to 17, or (4) change in VO_2_ less than 200 milliliters with an increase in work. Participants could stop the test at any point, and recovery involved walking at 2 mph and 0% grade for five minutes. The analyses were limited to participants who reached peak effort during graded exercise testing.

### 3.5 Statistical Analyses

All analyses were performed in R Statistical Software version 4.3.0 [31]. Demographic characteristics were compared between KL-VS_NC_ (N=93) and KL-VS_HET_ (N=43) using independent-sample t-tests for continuous measures and χ^2^ tests for categorical measures. To examine whether the relationship between VO_2peak_ and CSF biomarkers of core AD pathology, neurodegeneration, synaptic dysfunction, and neuroinflammation differed between KL-VS_NC_ and KL-VS_HET_, we fitted a series of linear regression models that incorporated a KL-VS* VO_2peak_ interaction term, while covarying for age, sex, age difference between CSF sampling and exercise test, *APOE* ε4 status, and parental history of AD. If the interaction was not significant, analyses were repeated after removing the KL-VS* VO_2peak_ term to examine the main effects of KL-VS and VO_2peak_ to assess whether CSF biomarker levels differed by either factor independently.

## 4 Results

### 4.1 Sample Characteristics

Table 1 details the background characteristics of the entire sample and by KL-VS genotype. The sample was predominantly white (98%) and female (64%), with an average age of 62.5±6.7 years and enriched for AD risk, with 38% carrying at least one *APOE* ε4 allele and 75% having a parental history of AD. VO_2peak_ averaged 26.0±6.4 mL/kg/min. For the entire sample, the average age difference between CSF sampling and the exercise test was 1.8±2.6 years. There were no significant differences in any of the above-mentioned characteristics between KL-VS_NC_ and KL-VS_HET_ (*P*s ≥ 0.17).

**Table 1.** Background characteristics of study participants.

|  | Entire Sample<br>(N = 136) | KL-VS<sub>NC</sub><br>(N = 93) | KL-VS<sub>HET</sub><br>(N = 43) | P |
| --- | --- | --- | --- | --- |
| **Age at CSF**<br>Mean (SD) | 62.5 (6.7) | 62.8<br>(6.7) | 62.4 (6.7) | 0.73 |
| **Female**<br>N (%) | 87 (64) | 61 (66) | 26 (61) | 0.83 |
| **White**<br>N (%) | 133 (98) | 91 (98) | 42 (98) | 0.38 |
| **APOE4+**<br>N (%) | 52 (38) | 36 (39) | 16 (37) | 0.78 |
| **Parental History of AD**<br>N (%) | 102 (75) | 71 (76) | 31 (72) | 0.78 |
| **Age<sub>exercise</sub> – Age<sub>LP</sub>**<br>Mean (SD) | 1.8 (2.6) | 1.5 (2.5) | 2.3 (2.9) | .17 |
| **VO<sub>2peak</sub> (mL/kg/min)**<br>Mean (SD) | 26.0 (6.4) | 26.2<br>(6.6) | 25.7 (5.9) | 0.57 |

\***Abbreviations:** AD = Alzheimer’s disease; Age<sub>LP</sub> = age at lumbar puncture; Age<sub>exercise</sub> = age at the exercise test visit; APOE4+ = carrier of at least one APOE ε4 allele; CRF = cardiorespiratory fitness; CSF = cerebrospinal fluid; KL- VS<sub>HET</sub> = *KLOTHO* KL-VS heterozygote; KL-VS<sub>NC</sub> = *KLOTHO* KL-VS non-carrier; mL/kg/min = milliliters per kilogram per minute; SD = standard deviation; VO<sub>2peak</sub> = peak oxygen consumption.

### 4.2 CSF biomarkers of core AD pathology as a function of the *KLOTHO* KL VS and VO_2peak_

A significant interaction was observed between KL-VS genotype and VO_2peak_ for pTau_181_ (*P=*0.03; Table 2). Figure 1 illustrates this relationship, whereby KL-VS_HET_ who were more fit had lower levels of pTau_181._ While KL-VSxVO_2peak_ interaction was not significant for Aβ_42_/Aβ_40,_ the levels differed by KL-VS genotype as indicated by a significant main effect of KL-VS (*P*=0.03). No significant interaction (*P*=0.36) nor main effects of either KL-VS or VO_2peak_ were observed for pTau_181_/Aβ_42_ (*Ps* ≥ 0.07).

**Figure 1.**
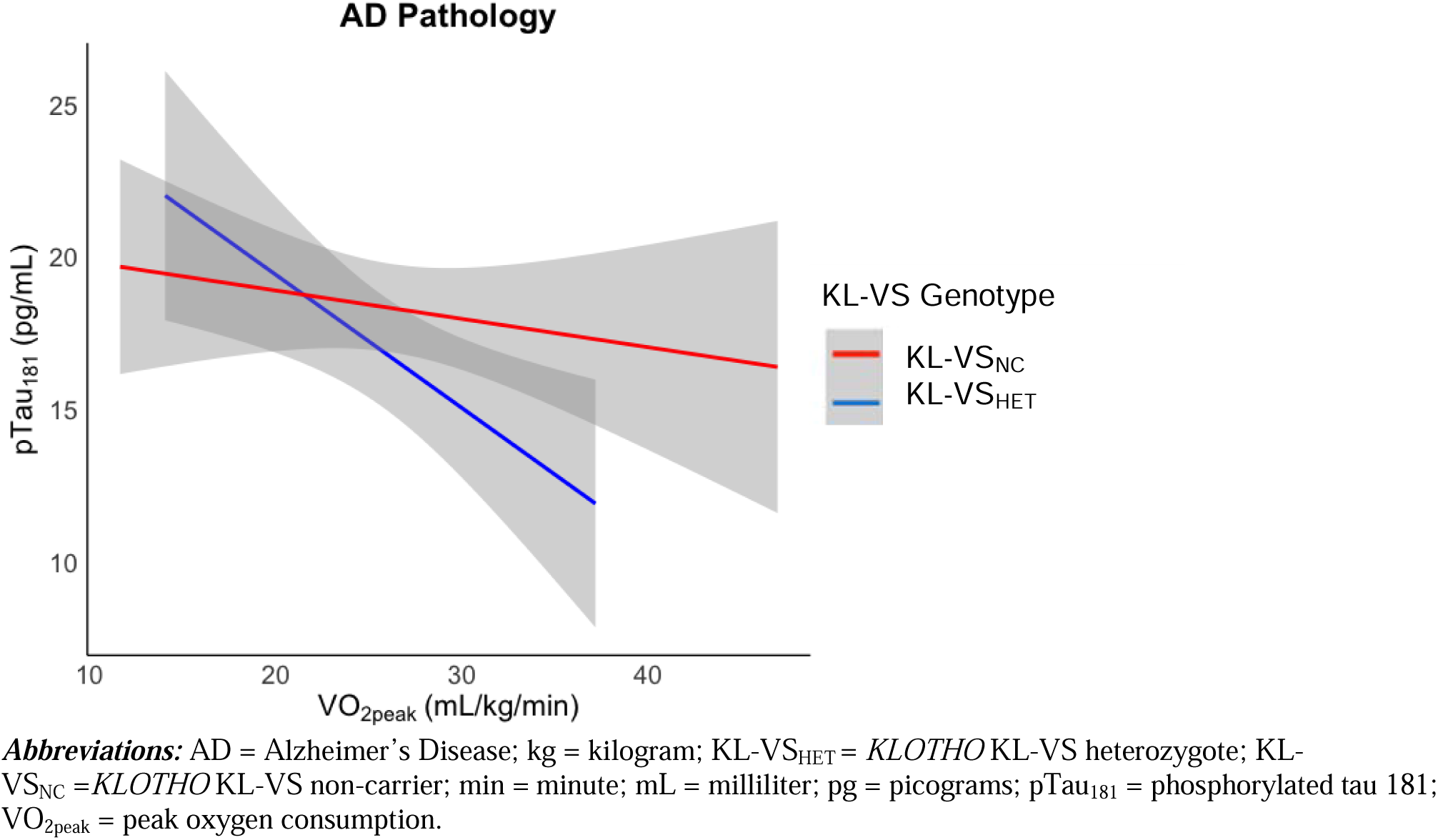
KL-VS genotype differences in CSF pTau_181_ based on CRF (VO_2peak_). KL-VS_HET_ with high CRF had lower levels of pTau_181_.

**Table 2.**
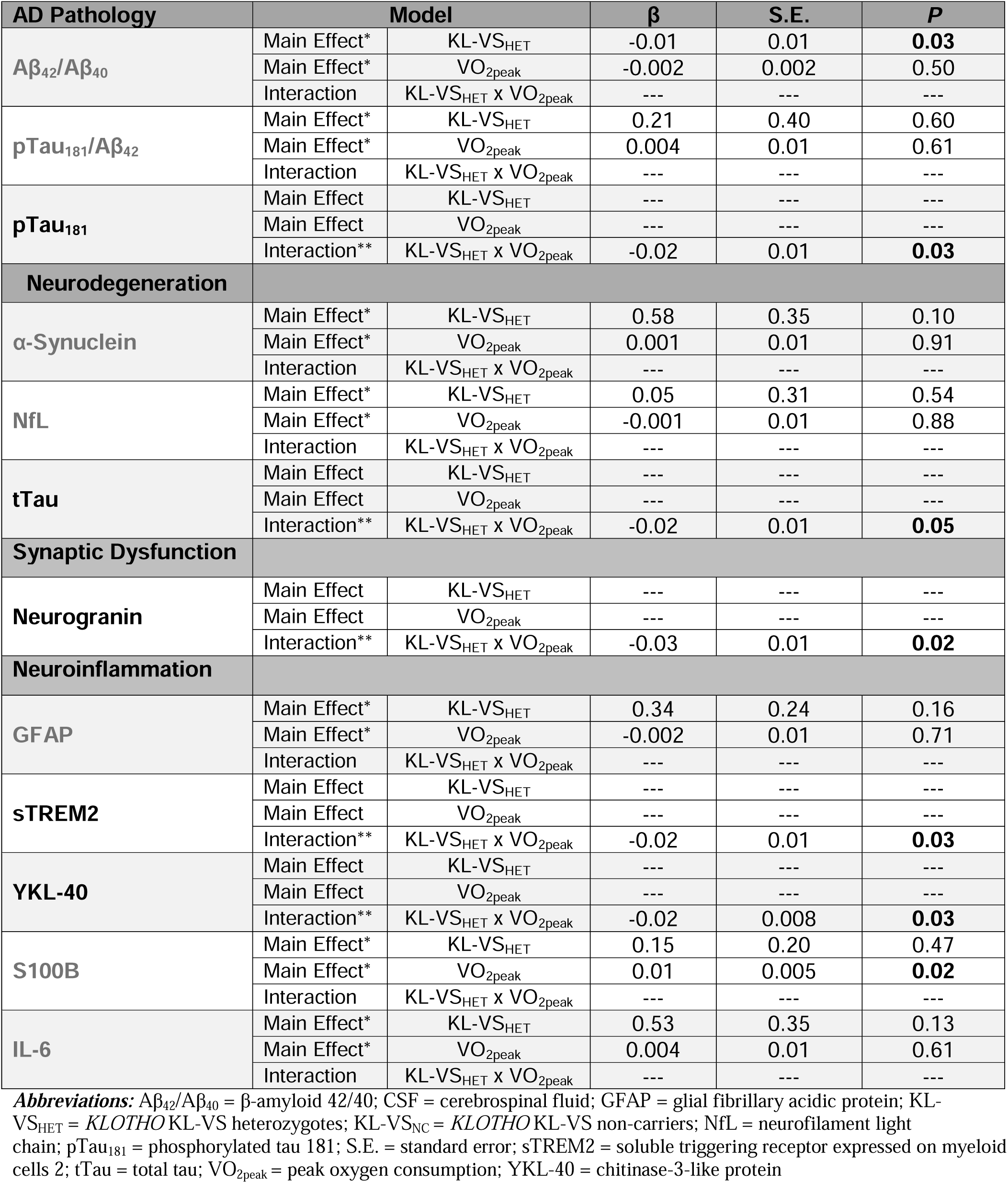
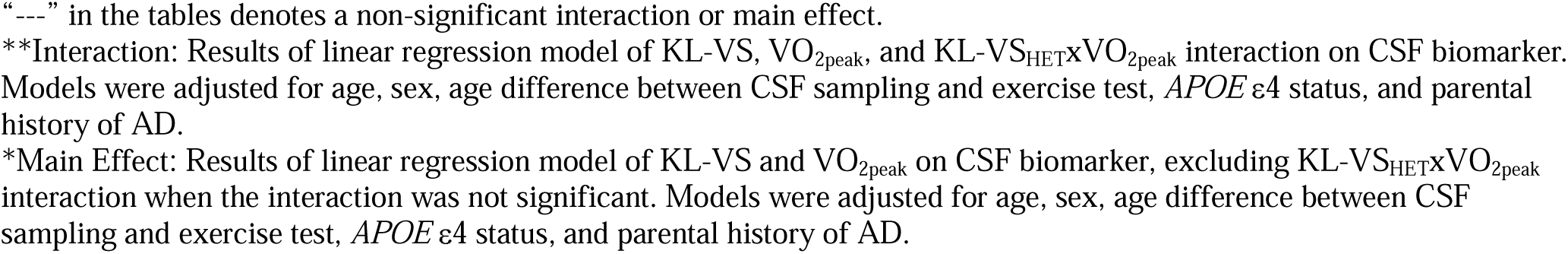
CSF biomarkers of core AD neuropathology, neurodegeneration, synaptic dysfunction, and neuroinflammation as a function of the *KLOTHO* KL VS and VO_2peak_.

| AD Pathology | | Model | $\beta$ | S.E. | P |
| --- | --- | --- | --- | --- | --- |
| Aβ<sub>42</sub>/Aβ<sub>40</sub> | Main Effect* | KL-VS<sub>HET</sub> | -0.01 | 0.01 | **0.03** |
|  | Main Effect* | VO<sub>2peak</sub> | -0.002 | 0.002 | 0.50 |
|  | Interaction | KL-VS<sub>HET</sub> x VO<sub>2peak</sub> | --- | --- | --- |
| pTau<sub>181</sub>/Aβ<sub>42</sub> | Main Effect* | KL-VS<sub>HET</sub> | 0.21 | 0.40 | 0.60 |
|  | Main Effect* | VO<sub>2peak</sub> | 0.004 | 0.01 | 0.61 |
|  | Interaction | KL-VS<sub>HET</sub> x VO<sub>2peak</sub> | --- | --- | --- |
| pTau<sub>181</sub> | Main Effect | KL-VS<sub>HET</sub> | --- | --- | --- |
|  | Main Effect | VO<sub>2peak</sub> | --- | --- | --- |
|  | Interaction** | KL-VS<sub>HET</sub> x VO<sub>2peak</sub> | -0.02 | 0.01 | **0.03** |
| Neurodegeneration |  |  |  |  |  |
| α-Synuclein | Main Effect* | KL-VS<sub>HET</sub> | 0.58 | 0.35 | 0.10 |
|  | Main Effect* | VO<sub>2peak</sub> | 0.001 | 0.01 | 0.91 |
|  | Interaction | KL-VS<sub>HET</sub> x VO<sub>2peak</sub> | --- | --- | --- |
| NfL | Main Effect* | KL-VS<sub>HET</sub> | 0.05 | 0.31 | 0.54 |
|  | Main Effect* | VO<sub>2peak</sub> | -0.001 | 0.01 | 0.88 |
|  | Interaction | KL-VS<sub>HET</sub> x VO<sub>2peak</sub> | --- | --- | --- |
| tTau | Main Effect | KL-VS<sub>HET</sub> | --- | --- | --- |
|  | Main Effect | VO<sub>2peak</sub> | --- | --- | --- |
|  | Interaction** | KL-VS<sub>HET</sub> x VO<sub>2peak</sub> | -0.02 | 0.01 | **0.05** |
| Synaptic Dysfunction |  |  |  |  |  |
| Neurogranin | Main Effect | KL-VS<sub>HET</sub> | --- | --- | --- |
|  | Main Effect | VO<sub>2peak</sub> | --- | --- | --- |
|  | Interaction** | KL-VS<sub>HET</sub> x VO<sub>2peak</sub> | -0.03 | 0.01 | **0.02** |
| Neuroinflammation |  |  |  |  |  |
| GFAP | Main Effect* | KL-VS<sub>HET</sub> | 0.34 | 0.24 | 0.16 |
|  | Main Effect* | VO<sub>2peak</sub> | -0.002 | 0.01 | 0.71 |
|  | Interaction | KL-VS<sub>HET</sub> x VO<sub>2peak</sub> | --- | --- | --- |
| sTREM2 | Main Effect | KL-VS<sub>HET</sub> | --- | --- | --- |
|  | Main Effect | VO<sub>2peak</sub> | --- | --- | --- |
|  | Interaction** | KL-VS<sub>HET</sub> x VO<sub>2peak</sub> | -0.02 | 0.01 | **0.03** |
| YKL-40 | Main Effect | KL-VS<sub>HET</sub> | --- | --- | --- |
|  | Main Effect | VO<sub>2peak</sub> | --- | --- | --- |
|  | Interaction** | KL-VS<sub>HET</sub> x VO<sub>2peak</sub> | -0.02 | 0.008 | **0.03** |
| S100B | Main Effect* | KL-VS<sub>HET</sub> | 0.15 | 0.20 | 0.47 |
|  | Main Effect* | VO<sub>2peak</sub> | 0.01 | 0.005 | **0.02** |
|  | Interaction | KL-VS<sub>HET</sub> x VO<sub>2peak</sub> | --- | --- | --- |
| IL-6 | Main Effect* | KL-VS<sub>HET</sub> | 0.53 | 0.35 | 0.13 |
|  | Main Effect* | VO<sub>2peak</sub> | 0.004 | 0.01 | 0.61 |
|  | Interaction | KL-VS<sub>HET</sub> x VO<sub>2peak</sub> | --- | --- | --- |

**Abbreviations:** Aβ <sub>42</sub>/Aβ <sub>40</sub> = β-amyloid 42/40; CSF = cerebrospinal fluid; GFAP = glial fibrillary acidic protein; KL-VS<sub>HET</sub> = *KLOTHO* KL-VS heterozygotes; KL-VS<sub>NC</sub> = *KLOTHO* KL-VS non-carriers; NfL = neurofilament light chain; pTau<sub>181</sub> = phosphorylated tau 181; S.E. = standard error; sTREM2 = soluble triggering receptor expressed on myeloid cells 2; tTau = total tau; VO<sub>2peak</sub> = peak oxygen consumption; YKL-40 = chitinase-3-like protein

### 4.3 CSF biomarkers of neurodegeneration, synaptic dysfunction and inflammation as a function of the *KLOTHO* KL VS and VO_2peak_

A significant KL-VSxVO_2peak_ interaction was observed for markers of neurodegeneration [**tTau**: *P*=0.05], synaptic dysfunction [**Ng**: *P*=0.02] and neuroinflammation [Table 2; **sTREM2**: *P*=0.03; **YKL-40**: *P*=0.03]. Figure 2A-D illustrates these findings; KL-VS_HET_ with greater VO_2peak_ had lower levels of tTau (A), Ng (B), YKL-40 (C) and sTREM2 (D). KL-VSxVO_2peak_ interaction was not significant for α-syn, NfL, GFAP, S100B or IL-6 (all *P*s>0.09). Additionally, no main effects of either KL-VS or VO_2peak_ were significant for α-syn, NfL, GFAP or IL-6 (all *P*s>0.10). S100B levels differed based on CRF, as indicated by a significant main effect of VO_2peak_ (*P*=0.02).

**Figure 2.**
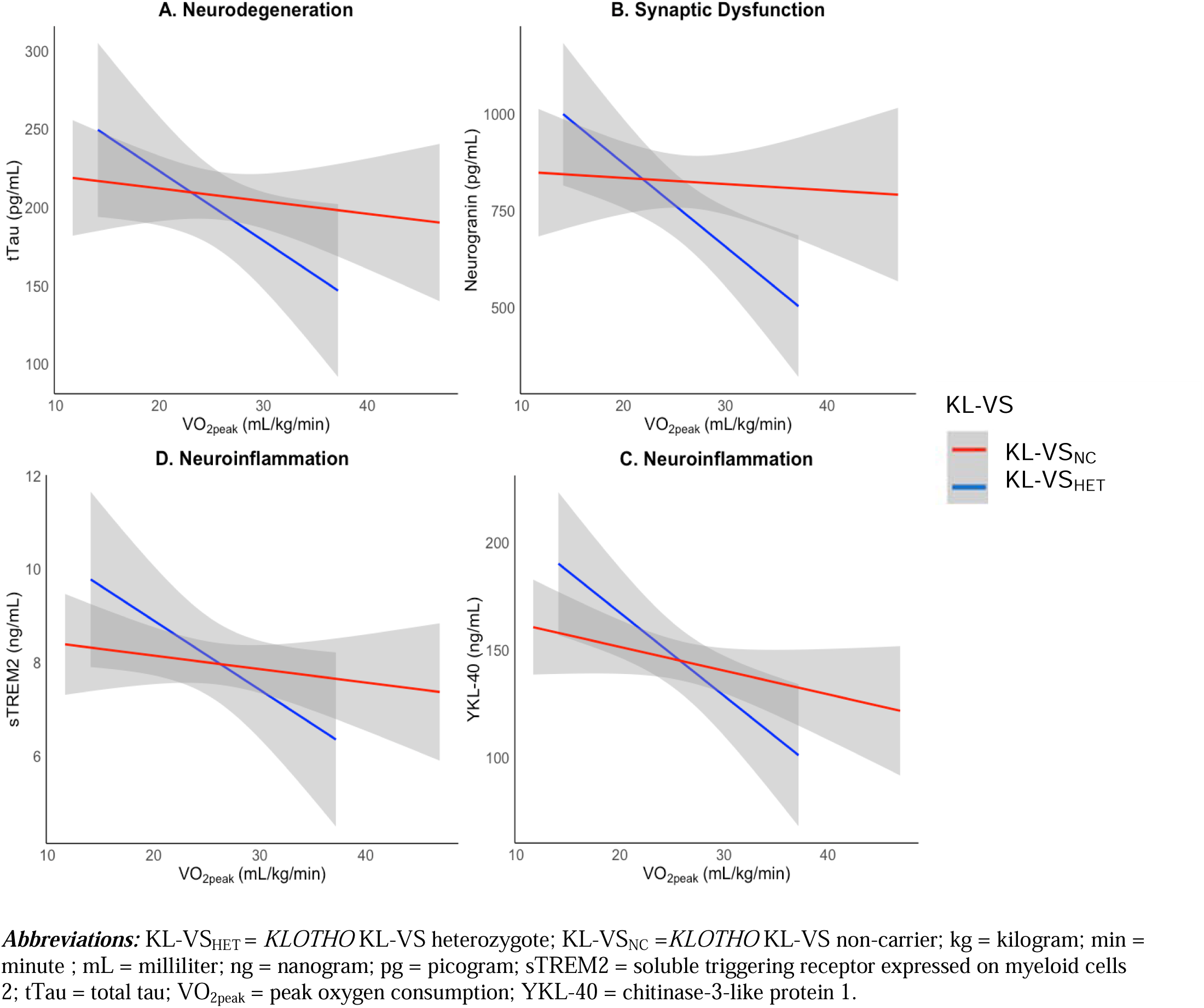
KL-VS differences in CSF A) tTau, B) Ng, C) YKL-40, and D) sTREM2 based on CRF (VO_2peak_). KL-VS_HET_ with high CRF had lower levels of neurodegeneration (tTau), synaptic dysfunction (Neurogranin) and neuroinflammation (sTREM2 and YKL-40).

## 5 Discussion

Our findings suggest that KL-VS_HET_ in conjunction with higher CRF may offer protection against a variety of adverse biomolecular changes associated with AD. Specifically, we report a synergistic relationship between KL-VS_HET_ and higher CRF with regard to core AD pathology (pTau_181_), neurodegeneration (tTau), synaptic dysfunction (Ng) and neuroinflammation (sTREM2 and YKL-40). Together, our results contribute to the growing literature supporting the role for *KLOTHO* in modulating age-related neuropathological processes and provide novel insights into how modifiable lifestyle factors, CRF specifically, may interact with genetic factors to influence AD risk.

The significant interaction between KL-VS_HET_ and CRF in relation to pTau_181_ and tTau suggests a key role for both fitness and genetic variation in mitigating tau-related pathology. Tau hyperphosphorylation and aggregation are hallmark features of AD, contributing to neuronal dysfunction and cognitive decline [27]. While CSF tTau and pTau are predictive markers of AD-related neurodegeneration and tangle formation, they are not direct markers of these processes [32]. pTau_181_ is also increasingly recognized as more indicative of Aβ-related tau dysmetabolism [32]. Furthermore, we caution against the simplistic interpretation of tTau as a marker of neurodegeneration given its nearly perfect correlation (∼98%) with pTau_181_ across our center-wide data [29], which suggests that the two measures may be largely reflective of overlapping rather than distinct pathological processes in our cognitively unimpaired cohort enriched for AD risk. Based on extant literature, we know that individuals with higher plasma tau who engage in more physical activity show a deceleration in cognitive decline [33]. Moreover, there is evidence that KL-VS_HET_ is associated with lower tau burden in aging adults [3, 4], suggesting that the KL-VS_HET_ genotype may play a role in tau clearance or stabilization. Our study advances this understanding by observing that, in KL-VS_HET_, higher CRF is associated with lesser CSF pTau and tTau levels, suggesting a combined role of genetic predisposition and physical activity in potentially slowing AD-related tau pathology. This supports the notion that physical activity may exert genotype-specific effects, providing greater neuroprotection in individuals with the KL-VS_HET_ genotype. Thus, increasing CRF through exercise may serve as a preventative therapeutic strategy for KL-VS_HET_ individuals, potentially mitigating AD-related tau pathology and its detrimental effects on cognitive health.

Ng is a postsynaptic protein expressed on postsynaptic spines of dendrites that is critical for synaptic plasticity and memory formation [34]. Elevated Ng levels in CSF are increasingly recognized as a marker of synaptic damage in AD [35]. Accumulating evidence suggests a protective effect of exercise on synaptic health [26, 36, 37]. For instance, recent literature showed that higher levels of physical activity are associated with lower Ng levels in individuals with low cardiovascular risk [36] and enhanced synaptic plasticity through the upregulation of brain-derived neurotrophic factor (BDNF) [37]. Animal models have demonstrated that elevated circulating protein klotho levels enhances synaptic plasticity and increases the expression of GluN2B, a key NMDA receptor subunit involved in synaptic transmission [16, 17]. Our findings add to this body of research, showing that those with higher CRF who concomitantly harbor the KL-VS_HET_ genotype have lower CSF Ng levels, indicative of preserved synaptic integrity. Furthermore, our results complement a recent study by our group, which reported that KL-VS_HET_ exhibit resilience against age-related increases in Ng levels [12]. While potentially protective effects of KL-VS_HET_ on synaptic health were present independent of fitness in our group’s previous work [12] our current findings suggest that higher CRF may amplify this neuroprotective effect, highlighting a potential synergistic protective relationship between KL-VS_HET_ and CRF against AD-related synaptic dysfunction.

Neuroinflammation plays a central role in the progression of AD, contributing to neuronal injury and the accumulation of amyloid and tau pathology [38, 39]. Two key biomarkers of neuroinflammation, sTREM2 and YKL-40, reflect activity in microglia and astrocytes, respectively [40–42]. sTREM2 is a soluble form of triggering receptor expressed in myeloid cells 2 (TREM2), which is predominantly expressed on microglia, the immune cells of the central nervous system [43]. Upon microglial activation, TREM2 signaling plays a crucial role in regulating microglial survival, proliferation, and phagocytic activity in response to neuronal injury or the accumulation of pathological proteins such as Aβ and tau. The release of sTREM2 into CSF reflects this activation process and is thought to act as a modulator of neuroinflammation, amplifying protective microglial responses [43]. In AD, the elevation of sTREM2 is seen across different stages of the disease, with the largest increase reported during the mild cognitive impairment (MCI) phase before transitioning to AD [44]. Moreover, CSF sTREM2 levels are significantly elevated in individuals with AD compared to healthy controls [45]. This suggests that microglial activation intensifies early in the disease process and persists throughout its progression, making sTREM2 a useful biomarker for tracking neuroinflammation within the context of neurodegeneration.

Similarly, YKL-40, a glycoprotein primarily expressed by astrocytes, acts as an early indicator of neuroinflammation and disease progression [42]. Growing evidence suggests that YKL-40 is also a promising biomarker for glial inflammatory response in AD, with significantly elevated CSF levels reported in individuals with AD compared to those cognitively unimpaired [42, 46], and also higher levels in *APOE* ε4 carriers with MCI [47]. Exercise modulates these neuroinflammatory markers in AD; CSF sTREM2 levels increase following physical activity, which may reflect transient microglial activation in response to acute exercise in individuals with AD [48]. However, it is important to note that individuals with AD typically exhibit elevated baseline CSF sTREM2 due to disease-related microglial activation [45], which may amplify the observed acute response. In contrast, our findings demonstrate that, in KL-VS_HET_, higher CRF is associated with lower CSF concentrations of both sTREM2 and YKL-40. This contrast suggests that while acute exercise may transiently activate microglia in individuals with AD, habitual exercise, reflected by higher CRF, may confer long-term neuroprotective effects by reducing chronic neuroinflammation in otherwise healthy populations. This implies a synergistic benefit of KL-VS_HET_ and physical fitness, suggesting that exercise could offer targeted benefits in reducing AD-related neuroinflammation in certain populations. Additionally, in the present study, higher CRF was associated with elevated S100B levels regardless of KL-VS genotype, contrary to what would be expected based on the literature. Although S100B is generally linked to inflammatory processes [49], its upregulation in individuals with higher CRF could indicate an adaptive response, promoting repair and neurotrophic effects in the brain rather than solely reflecting pathological inflammation. Further investigation is needed to clarify the role of S100B within the context of fitness and neurodegeneration. Overall, our findings reinforce the idea that klotho, which is inherently higher in KL-VS_HET_, and CRF might interact to mitigate neuroinflammatory processes in AD, providing a promising approach for slowing disease progression.

No significant interactions were observed for Aβ-related biomarkers. This is noteworthy given that the literature suggests that tau pathology might be more responsive to KL-VS heterozygosity compared to Aβ accumulation [4]. Moreover, KL-VS-related effects on Aβ are more pronounced in individuals carrying the *APOE* ε4 allele [13], potentially explaining the absence of significant findings related to Aβ in our sample. Further studies are needed to elucidate whether longitudinal fitness interventions yield stronger effects on these biomarkers in KL-VS_HET_, especially in *APOE* ε4-positive individuals. Together, the evidence underscores the complexity of genetic and environmental interplay in modulating AD-related biomolecular changes and emphasizes the importance of refining intervention strategies to target specific pathways and patient subgroups effectively [50].

This study is not without limitations. The cohort is predominantly white and well-educated, with a higher proportion of participants with parental history of AD or carrying at least one *APOE* ε4 allele compared to the general population. These characteristics may limit the generalizability of our results. Furthermore, as this is a cross-sectional study, we cannot assess the effects of the exposures on changes in CSF biomarker levels over time. Another potential limitation is the time lag between the lumbar puncture and the maximal graded exercise test; we did statistically adjust for this differential by incorporating the time lag as a covariate in the model. Notwithstanding the limitations, we believe this work makes a clinically meaningful contribution to the literature. Furthermore, WRAP and WADRC are ongoing longitudinal studies that continue to collect data and have also recently increased efforts to enroll participants from groups historically under-represented in research. Thus, future investigations will be able to examine how the interaction between KL-VS_HET_ and VO_2peak_ relates to prospective changes in the biomarkers examined in this study in larger, more diverse samples.

Overall, these findings suggest a positive synergy between KL-VS_HET_ and better CRF. While the precise mechanisms by which the KL-VS_HET_ genotype or CRF exerts protective effects are not entirely clear, a better understanding of lifestyle interventions aimed at improving fitness levels and genetic variants that modify AD risk hold promise to provide new avenues for prevention or treatment.

## Data Availability

All data produced in the present study are available upon reasonable request to the authors.

## Acknowledgements

We would like to acknowledge and thank the staff and study participants of the Wisconsin Registry for Alzheimer’s Prevention and the Wisconsin Alzheimer’s Disease Research Center and the laboratory technicians at the Clinical Neurochemistry Laboratory at the Mölndal campus, University of Gothenburg, Sweden, without whom this work would not be possible. This work was supported by National Institute on Aging grants R01AG077507 (O.C.O.), R01AG062167 (O.C.O.), R01AG085592 (O.C.O.), R01AG027161 (S.C.J), R01AG021155 (S.C.J.), R01AG054059 (C.E.G.) and P30AG062715 (S.A.); by National Institute of Neurological Disorders and Stroke grant R01NS092918 (D.B.D.); and a Clinical and Translational Science Award (UL1RR025011) to the University of Wisconsin, Madison. Portions of this research were supported by the Veterans Administration, including facilities and resources at the Geriatric Research Education and Clinical Center of the William S. Middleton Memorial Veterans Hospital, Madison, WI; European Research Council (#101053962); the Swedish Research Council (#2023-00356; #2022-01018 and #2019-02397); the Swedish Brain Foundation (#FO2017-0243); the Swedish Alzheimer Foundation (#AF-742881); the Swedish state under the agreement between the Swedish government and the County Councils, the ALF-agreement (#ALFGBG-715986; #ALFGBG-71320); and the Knut and Alice Wallenberg Foundation. H.Z. is a Wallenberg Scholar and a Distinguished Professor at the Swedish Research Council.

## Disclosures

Klotho is the subject of an international patent issued and held by the Regents of the University of California.

The NeuroToolKit is a panel of exploratory prototype assays designed to robustly evaluate biomarkers associated with key pathologic events characteristic of AD and other neurological disorders, used for research purposes only and not approved for clinical use (Roche Diagnostics International Ltd, Rotkreuz, Switzerland). Elecsys β-amyloid (1–42) and Elecsys Phospho-Tau (181P) CSF assays are approved for clinical use. COBAS and ELECSYS are trademarks of Roche. All other product names and trademarks are the property of their respective owners.

## References

1. 2024 Alzheimer’s disease facts and figures. Alzheimers Dement, 2024. 20(5): p. 3708–3821.

2. Van Cauwenberghe, C., C. Van Broeckhoven, and K. Sleegers, The genetic landscape of Alzheimer disease: clinical implications and perspectives. Genet Med, 2016. 18(5): p. 421–30.

3. Driscoll, I., et al., AD-associated CSF biomolecular changes are attenuated in KL-VS heterozygotes. Alzheimers Dement (Amst), 2022. 14(1): p. e12383.

4. Driscoll, I., et al., Age-Related Tau Burden and Cognitive Deficits Are Attenuated in KLOTHO KL-VS Heterozygotes. J Alzheimers Dis, 2021. 79(3): p. 1297–1305.

5. Boots, E.A., et al., Cardiorespiratory fitness is associated with brain structure, cognition, and mood in a middle-aged cohort at risk for Alzheimer’s disease. Brain Imaging Behav, 2015. 9(3): p. 639–49.

6. Vesperman, C.J., et al., Cardiorespiratory fitness and cognition in persons at risk for Alzheimer’s disease. Alzheimers Dement (Amst), 2022. 14(1): p. e12330.

7. Okonkwo, O.C., et al., Physical activity attenuates age-related biomarker alterations in preclinical AD. Neurology, 2014. 83(19): p. 1753–60.

8. Okonkwo, O. and H. van Praag, Exercise Effects on Cognitive Function in Humans. Brain Plast, 2019. 5(1): p. 1–2.

9. Gaitán, J.M., et al., Effects of Aerobic Exercise Training on Systemic Biomarkers and Cognition in Late Middle-Aged Adults at Risk for Alzheimer’s Disease. Front Endocrinol (Lausanne), 2021. 12: p. 660181.

10. Dougherty, R.J., et al., Cardiorespiratory fitness mitigates brain atrophy and cognitive decline in adults at risk for Alzheimer’s disease. Alzheimers Dement (Amst), 2021. 13(1): p. e12212.

11. Erickson, C.M., et al., KLOTHO heterozygosity attenuates APOE4-related amyloid burden in preclinical AD. Neurology, 2019. 92(16): p. e1878–e1889.

12. Driscoll, I.F., et al., KLOTHO KL-VS heterozygosity is associated with diminished age-related neuroinflammation, neurodegeneration, and synaptic dysfunction in older cognitively unimpaired adults. Alzheimers Dement, 2024. 20(8): p. 5347–5356.

13. Kurosu, H., et al., Suppression of aging in mice by the hormone Klotho. Science, 2005. 309(5742): p. 1829–33.

14. Buchanan, S., et al., Klotho, Aging, and the Failing Kidney. (1664-2392 (Print)).

15. Arking, D.E., et al., Association of human aging with a functional variant of klotho. Proc Natl Acad Sci U S A, 2002. 99(2): p. 856–61.

16. Dubal, D.B., et al., Life extension factor klotho enhances cognition. Cell Rep, 2014. 7(4): p. 1065–76.

17. Dubal, D.B., et al., Life extension factor klotho prevents mortality and enhances cognition in hAPP transgenic mice. J Neurosci, 2015. 35(6): p. 2358–71.

18. Belloy, M.E., et al., Association of Klotho-VS Heterozygosity With Risk of Alzheimer Disease in Individuals Who Carry APOE4. JAMA Neurol, 2020. 77(7): p. 849–862.

19. Liu, R., et al., Cardiorespiratory Fitness as a Predictor of Dementia Mortality in Men and Women. Medicine & Science in Sports & Exercise, 2012. 44(2): p. 253–259.

20. Vesperman, C.J., et al., Cardiorespiratory fitness attenuates age-associated aggregation of white matter hyperintensities in an at-risk cohort. Alzheimers Res Ther, 2018. 10(1): p. 97.

21. Dougherty, R.J., et al., Relationships between cardiorespiratory fitness, hippocampal volume, and episodic memory in a population at risk for Alzheimer’s disease. Brain Behav, 2017. 7(3): p. e00625.

22. Law, L.L., et al., Moderate intensity physical activity associates with CSF biomarkers in a cohort at risk for Alzheimer’s disease. Alzheimers Dement (Amst), 2018. 10: p. 188–195.

23. Liang, K.Y., et al., Exercise and Alzheimer’s disease biomarkers in cognitively normal older adults. Ann Neurol, 2010. 68(3): p. 311–8.

24. Brown, B.M., et al., Self-Reported Physical Activity is Associated with Tau Burden Measured by Positron Emission Tomography. J Alzheimers Dis, 2018. 63(4): p. 1299–1305.

25. Wang, M., et al., Exercise suppresses neuroinflammation for alleviating Alzheimer’s disease. J Neuroinflammation, 2023. 20(1): p. 76.

26. de Sousa Fernandes, M.S., et al., Effects of Physical Exercise on Neuroplasticity and Brain Function: A Systematic Review in Human and Animal Studies. Neural Plast, 2020. 2020: p. 8856621.

27. Masters, C.L., et al., Alzheimer’s disease. Nat Rev Dis Primers, 2015. 1: p. 15056.

28. Johnson, S.C., et al., The Wisconsin Registry for Alzheimer’s Prevention: A review of findings and current directions. Alzheimers Dement (Amst), 2018. 10: p. 130–142.

29. Van Hulle, C., et al., An examination of a novel multipanel of CSF biomarkers in the Alzheimer’s disease clinical and pathological continuum. Alzheimers Dement, 2021. 17(3): p. 431–445.

30. Medicine, A.C.o.S., ACSM’s guidelines for exercise testing and prescription. 2013: Lippincott williams & wilkins.

31. R Development Core Team, R: A language and environment for statistical computing. 2021, R Foundation for Statistical Computing: Vienna, Austria.

32. Zetterberg, H. and K. Blennow, Moving fluid biomarkers for Alzheimer’s disease from research tools to routine clinical diagnostics. Mol Neurodegener, 2021. 16(1): p. 10.

33. Desai, P., et al., Longitudinal Association of Total Tau Concentrations and Physical Activity With Cognitive Decline in a Population Sample. JAMA Netw Open, 2021. 4(8): p. e2120398.

34. Gerendasy, D.D. and J.G. Sutcliffe, RC3/neurogranin, a postsynaptic calpacitin for setting the response threshold to calcium influxes. Mol Neurobiol, 1997. 15(2): p. 131–63.

35. Kester, M.I., et al., Neurogranin as a Cerebrospinal Fluid Biomarker for Synaptic Loss in Symptomatic Alzheimer Disease. JAMA Neurol, 2015. 72(11): p. 1275–80.

36. Stojanovic, M., et al., Effect of exercise engagement and cardiovascular risk on neuronal injury. Alzheimers Dement, 2023. 19(10): p. 4454–4462.

37. Lu, Y., et al., Recent advances on the molecular mechanisms of exercise-induced improvements of cognitive dysfunction. Transl Neurodegener, 2023. 12(1): p. 9.

38. Kinney, J.W., et al., Inflammation as a central mechanism in Alzheimer’s disease. Alzheimers Dement (N Y), 2018. 4: p. 575–590.

39. Leng, F. and P. Edison, Neuroinflammation and microglial activation in Alzheimer disease: where do we go from here? Nat Rev Neurol, 2021. 17(3): p. 157–172.

40. Suárez-Calvet, M., et al., Early increase of CSF sTREM2 in Alzheimer’s disease is associated with tau related-neurodegeneration but not with amyloid-β pathology. Mol Neurodegener, 2019. 14(1): p. 1.

41. Nordengen, K., et al., Glial activation and inflammation along the Alzheimer’s disease continuum. Journal of Neuroinflammation, 2019. 16(1): p. 46.

42. Connolly, K., et al., Potential role of chitinase-3-like protein 1 (CHI3L1/YKL-40) in neurodegeneration and Alzheimer’s disease. Alzheimers Dement, 2023. 19(1): p. 9–24.

43. Suárez-Calvet, M., et al., sTREM2 cerebrospinal fluid levels are a potential biomarker for microglia activity in early-stage Alzheimer’s disease and associate with neuronal injury markers. EMBO Mol Med, 2016. 8(5): p. 466–76.

44. Hok-A-Hin, Y.S., et al., Neuroinflammatory CSF biomarkers MIF, sTREM1, and sTREM2 show dynamic expression profiles in Alzheimer’s disease. Journal of Neuroinflammation, 2023. 20(1): p. 107.

45. Heslegrave, A., et al., Increased cerebrospinal fluid soluble TREM2 concentration in Alzheimer’s disease. Mol Neurodegener, 2016. 11: p. 3.

46. Mavroudis, I., et al., YKL-40 as a Potential Biomarker for the Differential Diagnosis of Alzheimer’s Disease. Medicina (Kaunas), 2021. 58(1).

47. Wang, L., et al., Cerebrospinal fluid levels of YKL-40 in prodromal Alzheimer’s disease. Neurosci Lett, 2020. 715: p. 134658.

48. Jensen, C.S., et al., Exercise as a potential modulator of inflammation in patients with Alzheimer’s disease measured in cerebrospinal fluid and plasma. Exp Gerontol, 2019. 121: p. 91–98.

49. Donato, R., S100: a multigenic family of calcium-modulated proteins of the EF-hand type with intracellular and extracellular functional roles. Int J Biochem Cell Biol, 2001. 33(7): p. 637–68.

50. Sperling, R.A., C.R. Jack, Jr., and P.S. Aisen, Testing the right target and right drug at the right stage. Sci Transl Med, 2011. 3(111): p. 111cm33.

